# N-acetylcysteine to reduce kidney and liver injury associated with drug-resistant tuberculosis treatment

**DOI:** 10.1101/2025.01.15.25320552

**Authors:** Idu Meadows, Happiness Mvungi, Kassim Salim, Oscar Kaswaga, Peter Mbelele, Alphonce Liyoyo, Hadija Semvua, Athumani Ngoma, Scott Heysell, Stellah Mpagama

## Abstract

**Background:** New drug classes and regimens have shortened the treatment duration for drug-resistant tuberculosis, but adverse events (AEs) and organ toxicity remain unacceptably common. N-acetylcysteine (NAC) has demonstrated potential in reducing kidney and liver toxicity in other clinical settings, but efficacy in drug-resistant tuberculosis treatment has not been rigorously evaluated.

**Method:** A randomized controlled trial (PACTR202007736854169) was conducted at Kibong’oto Infectious Disease Hospital in Tanzania to assess the efficacy of NAC in reducing AEs in patients undergoing rifampin-resistant pulmonary tuberculosis treatment. Participants received an all-oral standardized rifampin-resistant regimen alone, with NAC 900 mg daily, or NAC 900 mg twice daily for 6 months. AEs, severe AEs, and renal and liver toxicity were monitored monthly and classified according to the Risk, Injury, Failure, Loss, and End-stage kidney disease criteria and National Cancer Institute Common Terminology Criteria for Adverse Events. Incident ratios and Kaplan-Meier curves were employed to compare group event occurrences.

**Results:** 66 patients (mean age 47±12 years; 80% male) were randomized into three groups of 22. One hundred and fifty-eight AEs were recorded: 52 (33%) in the standard treatment group, 55 (35%) in the NAC 900 mg daily group, and 51 (32%) in the NAC 900 mg twice daily group (p>0.99). Severe AEs were observed in 4 patients in the standard group, 2 in the NAC 900 mg daily group, and 3 in the NAC 900 mg twice daily group. Renal toxicity was more prevalent in the standard treatment group (45% vs. 23%; p=0.058), with a shorter onset of time to toxicity (*χ*2 = 3.199; p=0.074). Liver injury events were rare across all groups.

**Conclusion:** Among Tanzanian adults receiving rifampin-resistant tuberculosis treatment, NAC did not significantly reduce overall AEs but demonstrated important trends in reducing renal toxicity.

## Introduction

Tuberculosis (TB) diseases remain a global concern, with 10.6 million people affected by the disease in the 2023 Global TB Report.^1^ Of these, 410,000 developed drug-resistant tuberculosis (DR)-TB, and 1.3 million total deaths were reported, with 160,000 attributed to DR-TB. DR-TB is classified into two categories: rifampin-resistant or multi-drug-resistant (RR/MDR) and extensively drug-resistant (XDR) based on the approach to treating these resistance patterns. RR/MDR-TB is resistant to at least isoniazid and rifampin, the cornerstone medicines for the treatment of otherwise susceptible TB.^2^ XDR-TB fulfills the definition of MDR/RR-TB resistance with resistance to anti-TB fluoroquinolones and at least one additional group A drug (group A drugs are the most potent among those typically used for drug-resistant forms of TB using more extended treatment regimens and comprise levofloxacin, moxifloxacin, bedaquiline, and linezolid).^1–4^ Earlier, second-line DR-TB treatments included 20-24 months of drugs that were deemed less effective and more toxic.^5,6^ Recent advancements, as seen in such trials as TB-PRACTECAL and ZeNix, have led to more efficacious and shorter-duration treatments. Currently, numerous trials and operational research are testing combinations of group A drugs and other recently introduced agents in varying durations for DR-TB.^4^

While an intense focus has been placed on improving efficacy and treatment duration, toxicity and intolerability remain a significant barrier to treatment success.^7–9^ Adverse events are unexpected negative responses to a medicine or vaccine exposure.^7^ The proportion of people experiencing estimated AEs associated with drug-susceptible TB treatment has varied widely (8%-85%). Still, it is more consistently reported among people treated with drugs used in DR-TB regimens (69%-96%).^6^ In East Africa, AEs reported among Uganda, Ethiopia, and Tanzania were 67.4% of 120 patients^6^, 98.6% of 72,^6,10^, with 87.7%^11^ of 260 patients analyzed, respectively. AEs can result in interruptions, treatment failures, acquired resistance, prolonged hospital stays, or death in DR-TB patients.^12^ AE related to DR-TB treatments can affect different organ systems but commonly affect the kidney and liver.^13–15^

N-acetylcysteine (NAC) has emerged as an AE prevention therapy in some drug-related organ injuries due to its anti-oxidative and anti-inflammatory actions. NAC has two ways to counteract oxidative stress: indirect and direct anti-oxidative activity. In indirect anti-oxidative activity, NAC is a thiol that acts as an acetylated precursor to the amino acid L-cysteine, contributing to glutathione production and countering the oxidative stress associated with glutathione depletion. Through the NAC-free thiol group, it binds with active redox metal ions and can react with reactive oxygen and nitrogen species in direct antioxidant activity.^16–18^

NAC’s anti-inflammatory activity also originates from inhibiting the pro-NF-kappa beta pathway, suppressing inflammatory cytokine release.^17–20^ NAC is metabolized through the liver and excreted via the renal system. In healthy adults, its life span is 5.6 hours.^16,21^ NAC has been used to decrease the risk of contrast-induced renal toxicity in patients with myocardial infarction undergoing primary percutaneous coronary intervention.^22,23^

Most studies have employed NAC for liver injury prevention in drug-susceptible TB treatment, acetaminophen overdose, or alcohol-induced liver injury.^12,16–19,24–26^ However, little is known about NAC’s protective effects against AE in DR-TB treatment. Therefore, a randomized control trial was conducted among adults initiating pulmonary RR/MDR-TB treatment at Kibong’oto Infectious Disease Hospital, Tanzania (PACTR202007736854169) registered 03 July 2020. The primary objective was to assess the occurrence of clinical or laboratory-based AE during the initial six months of therapy in patients utilizing a standard treatment group, NAC of 900 mg oral daily, or NAC of 900 mg oral twice daily, in conjunction with a standardized all-oral, bedaquiline based RR/MDR-TB regimen.^12^

## Materials and Methods

The inclusion criteria were age range from 18-75 years, newly diagnosed RR or MDR-TB without additional fluoroquinolone or XDR-TB resistance pattern, no prior diagnosis or RR/MDR-TB treatment, a Karnofsky score of 50 or above defined as individuals requiring less considerable or frequent medical care, female participants had a negative urinary pregnancy test, kidney function with creatinine < 2mg/dL creatinine or creatinine clearance level >30 ml/min per Cockcroft-gault formula, hemoglobin > or 8 g/dL, platelets > 100k/mm3, AST/ALT or bilirubin 2X upper limit normal (ULN), and an electrocardiogram (EKG) with normal sinus rhythm at baseline. Patients were excluded if they had a known allergy or hypersensitivity to NAC, were living with HIV that met the WHO clinical stage 4 disease criteria or had CD4 count < 100 cells/mm, had a, history of cardiac arrhythmias or QTc prolongation, were pregnant, exposed to IV fungal medication within the last 90 days, had previous or existing brain pathology (major head trauma, meningitis, encephalitis, metastasis, vestibular schwannoma), or had participated in other clinical trials with investigational agents within the prior eight week.

### Ethical considerations

Eligible participants were given a detailed review of the study’s specifics, and the study staff discussed and answered any concerns or questions. Participants also gave written informed consent for the study, which was approved by the Tanzanian National Institute of Medical Research.

### Consent for publications

The details of any images in this study can be published. All authors providing consent to the publication of the images have been shown the article contents to be published.

### Interventions

NAC 900 mg was procured in tablet form and manufactured by BioAdventex Pharma Inc. Randomization groups included the standardized regimen group, NAC 900 mg orally once daily, or NAC 900 mg orally twice daily with standardized regimen treatment. The standardized treatment background regimen of rifampin resistance (RR)/MDR-TB treatment. The standardized regimen, termed RISE-**R**emoved **I**njectable **S**hort-Course for ***e***Xpert-diagnosed RR/MDR-TB, was an all-oral regimen constructed from regional *M. M.tuberculosis* drug-susceptibility patterns and studied operationally at multiple sites in Tanzania.^27^ Daily dosed bedaquiline was given for the first 6 months, linezolid for the first 2 months, and levofloxacin, clofazimine, pyrazinamide, and cycloserine for the entire duration of 9 months and extended up to 12 months if delayed clearance of *M. tuberculosis* from the sputum. Delaminid was used as an alternative if known resistance to one drug in the regimen or drug-specific intolerance developed. The Hain Genotype MTBs line probe assay was available for testing for common *gyrA* mutations conferring fluoroquinolone resistance.

AE severity was graded according to Common Terminology Criteria for AE (CTCAE) version 5.0 from the U.S. Department of Health and Human Services National Cancer Institute, published in November 2017. Renal toxicity was graded with Risk Injury Failure, Loss and End-stage Kidney disease (RIFLE) for early identification of renal toxicity.

Participants were actively followed weekly from treatment initiation through week 24 for symptom screen and targeted physical exam for evaluation. Scheduled complete blood count and comprehensive metabolic panel were performed at treatment initiation (baseline), week two, week four, and every four weeks through week 24. EKGs were obtained at baseline, week 1, week 2, week 4, and then every four weeks through week 24. Chest X-ray was obtained at baseline and week 24. Per routine clinical practice, sputum was collected at baseline and weekly until the Acid-fast bacilli (AFB) smear and mycobacterial culture were negative.

### Outcome measures

The primary outcome measure was the number of AE’s and total number of severe AEs (SAEs) in each group through 26 weeks, two weeks after the final study medication doses. The secondary outcomes, including the total number and time to events of renal toxicity, liver toxicity, and anemia, were chosen based on the expected higher frequency and prior evidence from other settings that NAC may prevent or delay drug-induced renal and liver toxicity while there was not expected impact on anemia. These measures serve as an internal control check against a spurious finding.

### Statistical analysis

The primary outcome was comparing the incidence of total AEs and SAEs in each arm, which was calculated using counts and proportions. Differences in the incidence of total AEs and SAEs and individual categories of AEs among each group were analyzed using Pearson Chi-square and Fisher Exact test. For secondary renal and liver injury and anemia outcomes, both NAC treatment groups were combined and compared to the standard treatment group with Kaplan-Meier estimates with 95 % CIs plotted for time-to-event analysis. The statistical analyses were performed using IBM SPSS Statistics for Windows, Version 29. *P* values of <0.05 were considered statistically significant.

### Metadata will be available in repositories

## Results

From Sept 18th, 2020, to October 27th, 2022, 70 patients were screened, and 66 met the eligibility criteria (Figure 1). Sixty-six were randomized to standard background RR-TB treatment (n=22), standard RR-TB treatment with NAC 900 mg daily (n=22), or NAC 900 mg twice daily (n=22) (Figure 1). The average age was 47 with a standard deviation (SD) ± 12. There were no differences in baseline creatinine, AST or ALT, and hemoglobin among the randomization groups (Table 1). Other demographic characteristics are presented in Table 1. Seven patients (10.6%) withdrew from the study, 6 voluntarily and 1 due to pregnancy. Two additional patients died during the study, 1 patient in the NAC daily group 1 patient in the NAC twice daily group.

**Figure 1:**
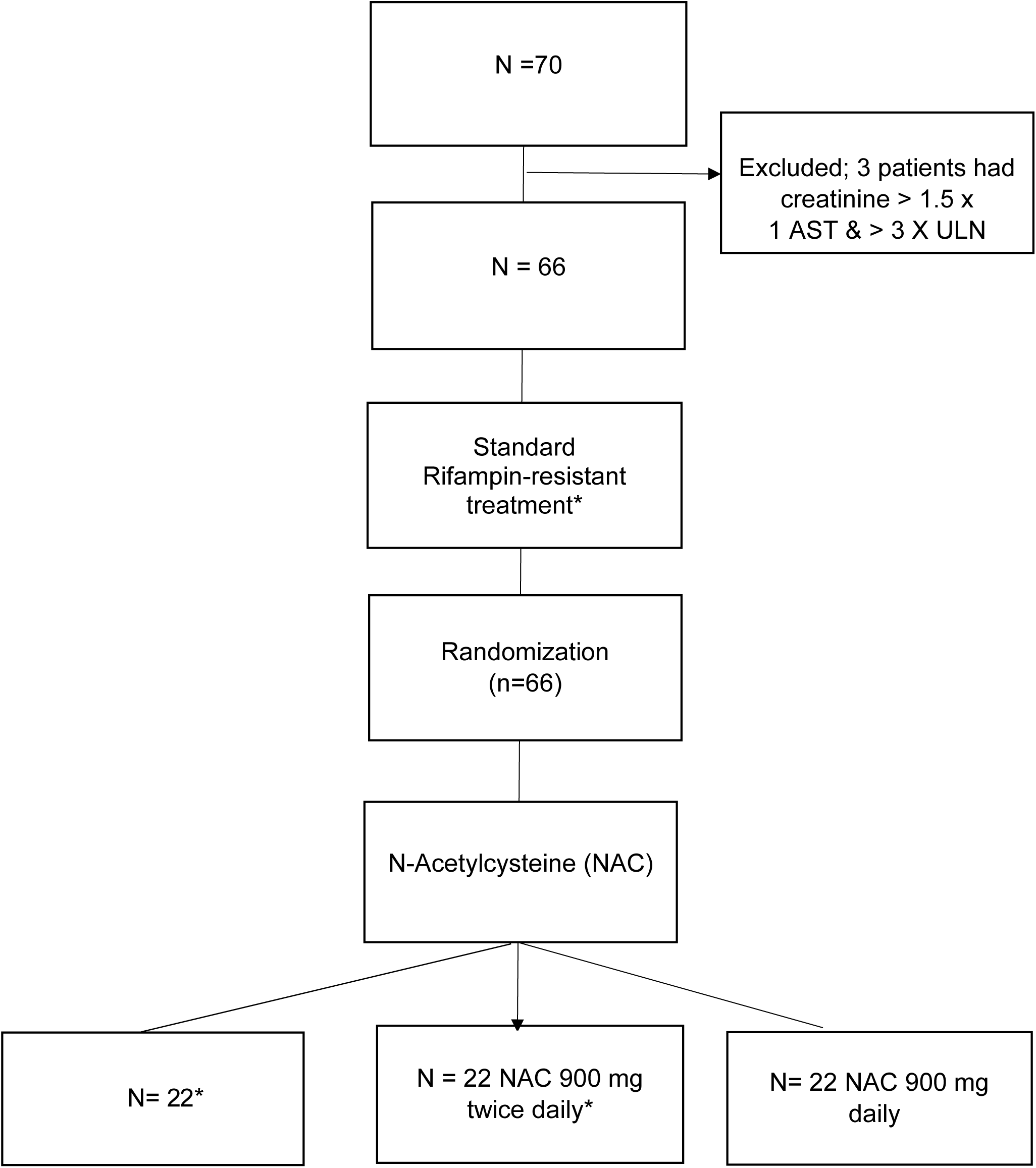
study flow chart displaying enrollment and randomization of study participants. *Bedaquiline, Levofloxacin, cysloserine, clofazimine, pyrazinamide, Linezolid, alternative: Delaminid. aspartate aminotransferase (AST), Alanine aminotransferase (ALT), upper limit normal (ULN)

**Table 1.**
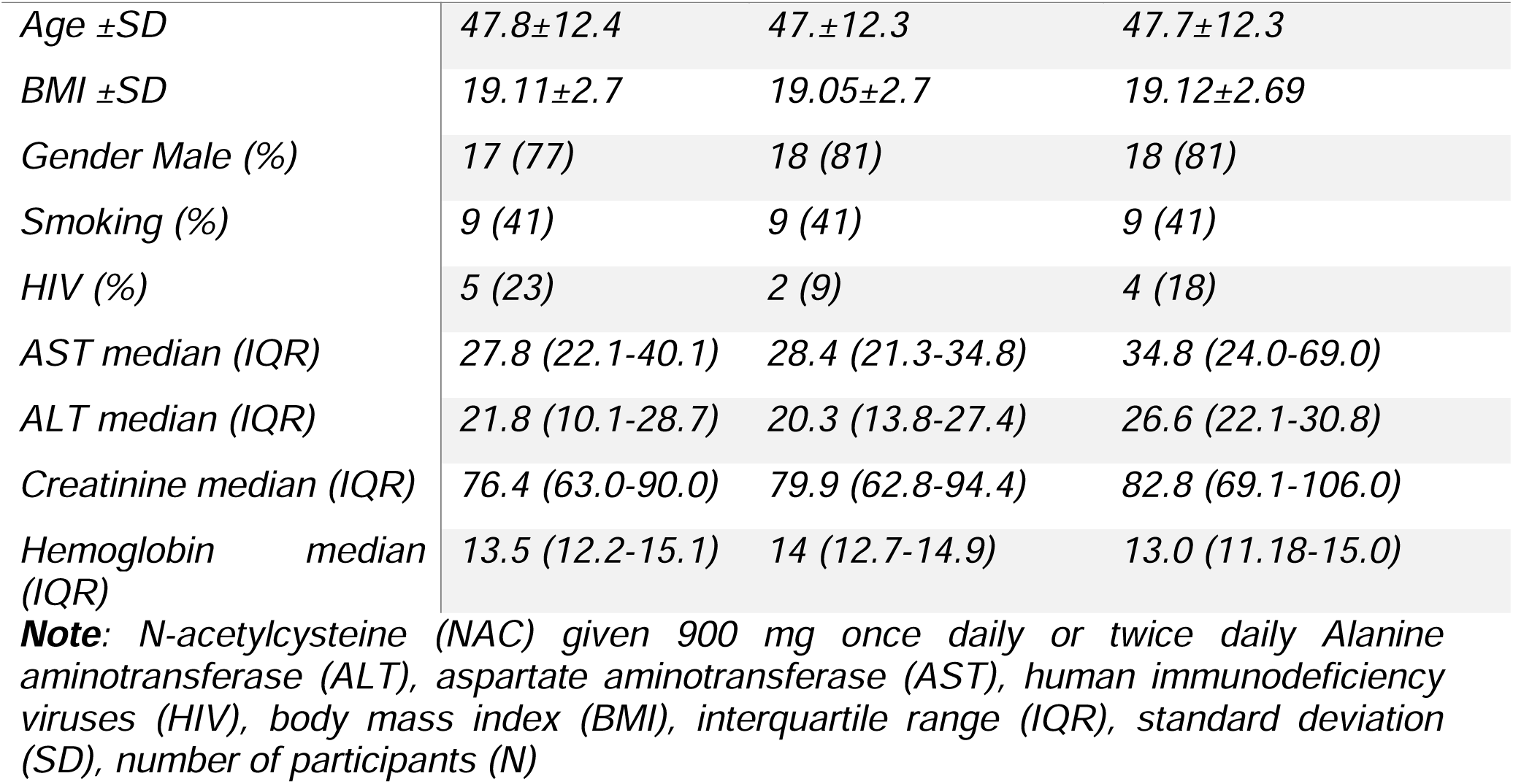
Demographics and baseline laboratory results of study participants (N = 66)

**Table 2.**
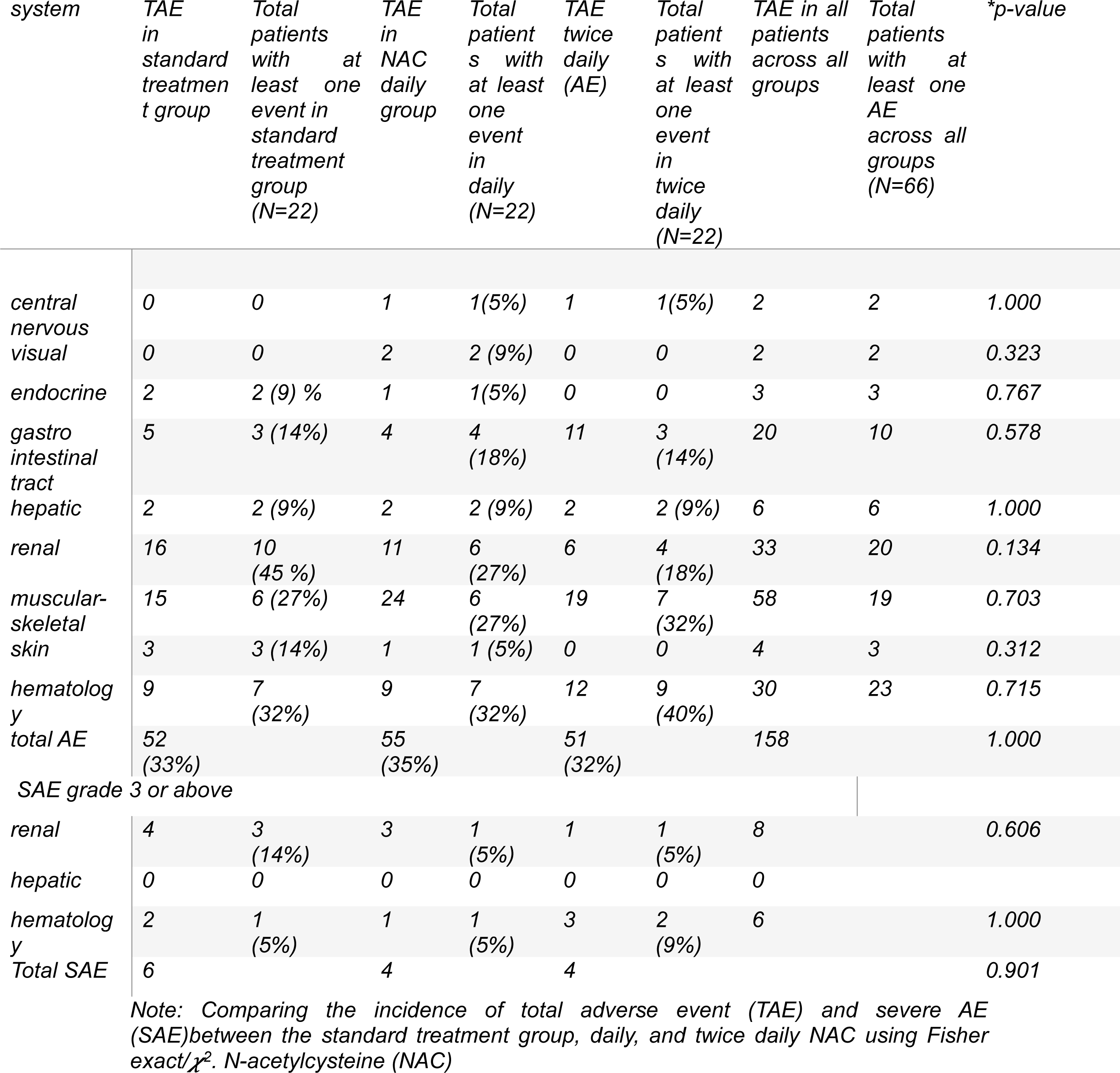
Incidence of AE in interventional group vs standard treatment group of (N= 66) participants.

### Incidence of adverse events

There were 158 total AEs, with 33 % in standard treatment, 35 % in daily NAC, and 32% in twice-daily NAC groups (P>0.99). The renal toxicities in the standard treatment group were 45% (10 of 22) compared to daily NAC with 27% (6 of 22) and twice daily NAC with 18% (4 of 22). When both NAC groups were combined, 23% (10 of 44) experienced renal toxicity compared with the standard treatment group Supplemental Table 1. Liver toxicity occurred in 9% (2 of 22) in each group and anemia in 32% (7 of 22) in the control and daily NAC group, and 36 % (16 of 44) in NAC group.

The total SAEs that occurred were 42.9 % (6 of 14) in the standard treatment group and 29% (4 of 14) in each NAC group (*P*=0.901). Severe renal toxicity occurred in 18% (4 of 22) of the standard treatment group and only 5% (1 of 22) in each NAC group. There was no severe liver injury in any of the groups. Severe anemia occurred in 5% (1 of 22) in both the control and daily NAC group and 9% (2 of 22) twice daily NAC.

### Time to individual adverse events

A log-rank test was run to determine if there were differences in the time to event of the pre-specified secondary outcome of interests (renal toxicity, liver toxicity, anemia) among those that received NAC (combined NAC daily and NAC twice daily groups). The time to renal toxicity trended toward a shorter time for those not receiving NAC, *χ* (1) = 3.199, p = .074 (Figure 2). There was no difference in time to events for Liver toxicity or anemia (supplemental figures S1 & S2)

**Figure 2:**
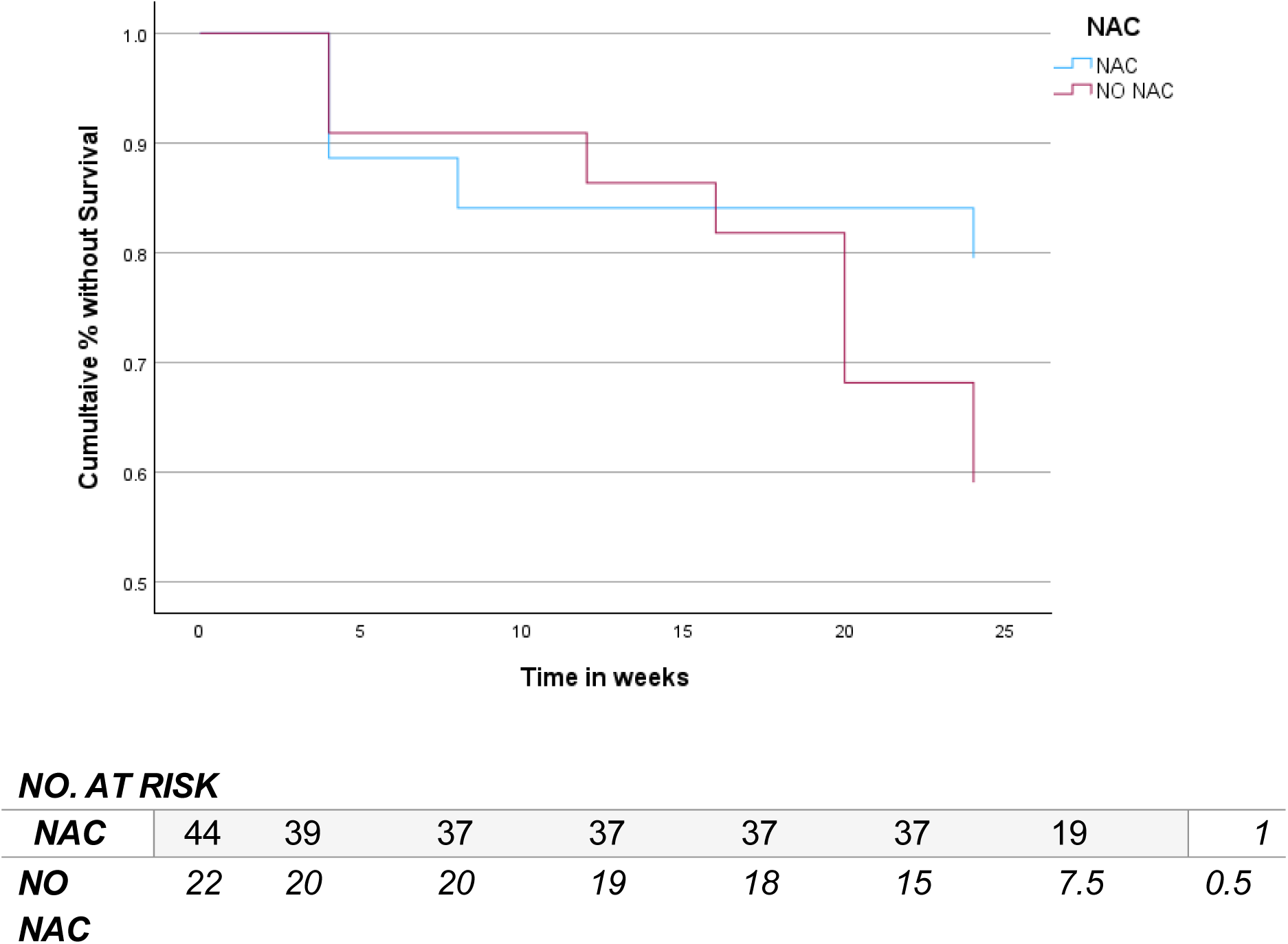
Time to event curve of renal injury through 24 weeks of standard treatment and NAC group. Note: N-acetylcysteine (NAC)

## Discussions

In this phase 2b randomized controlled trial among adults starting RR/MDR-TB treatment once or twice daily, NAC did not significantly reduce the total number of AEs or SAEs over the first 26 weeks of treatment compared to placebo. However, important trends suggest that NAC may reduce the number and severity of renal toxicity events. There was no difference in the total number or time for the development of liver injury among the treatment groups. Yet, the overall incidence of liver injury in the study population was lower than expected.

NAC has been found to protect against acute renal toxicity in high-risk populations with diabetes, cardiac disease, chronic kidney disease, or other chronic illnesses during events such as contrast exposure, coronary artery bypass surgery, and kidney transplantation have been discussed.^22,23,28–30^ Most explanations of NAC’s effects on renal toxicity suggest that it prevents renal vasoconstriction, inflammation, and oxidative stress. Increasing endothelial nitric oxide synthase expression, nitric oxide, and prostaglandin E2 production and reducing angiotensin-converting enzyme activities, thus preventing renal constriction.^22,31,32^ It reduces oxidative stress related to ischemic reperfusion or drugs by eliminating free radicals and indirectly generating glutathione. Additionally, NAC may down-regulate inflammation by inhibiting the transcription of activator protein one and nuclear factor kappa-light-chain-enhancer and reducing lymphocyte and macrophage infiltrations.^21,23,28–31,33^ Thus, the putative activity of NAC is more likely to prevent toxicity that could occur with multiple drugs over a long period, such as with RR/MDR-TB treatment. As observed in our study, NAC appeared to prevent more late-onset kidney toxicity.

Interestingly, the incidence of renal toxicity in our study was 30 % of 66 participants, unlike other studies that have reported fewer participants experiencing renal toxicity with similar drug regimens. For instance, the ZeNix trial reported 13.3 % renal toxicity in 45 participants who received bedaquiline, pretomanid, and linezolid 600 mg for 26 weeks.^9^ In the TB Practecal trial, 2.8 % of 142 participants experienced renal toxicity during treatment with a 24-week regimen of bedaquiline, pretomanid, linezolid, and moxifloxacin.^34^ The variation in renal toxicity incidence may be explained by factors such as baseline or reference serum creatinine, the inclusion/exclusion criteria, and the difference in renal toxicity diagnosis criteria used in each study.^35^ For instance, these studies used the Division of Microbiology and Infectious Disease Table to grade renal toxicity severity (DMIDS); in contrast, we used RIFLE criteria, a more pragmatic approach for early identification of participants at risk of developing renal toxicity risk than those of other studies, which likely led to the classification of more patients with renal toxicity in our study.^36,37^ Another explanation was the difference in the median age of our study was 47 compared to both studies, ZeNix and Practecal trials, where the median age was 34 and 38, respectively, and increased age is related to decreased drug clearance rate, thereby increasing the risk of renal toxicity in our study.^10^ Another explanation is the difference in drug regimens used, where our study included pyrazinamide and cycloserine, which both have been reported to cause renal toxicity.^38^

Other uses of NAC as an adjunctive therapy in TB include its use in the prevention of drug-induced liver injury in drug-susceptible TB treatment, where first-line drugs with significant liver injury profiles are used in combination (isoniazid, rifampin, and pyrazinamide). The hepatoprotective effect is mainly related to NAC’s promoting glutathione regeneration, detoxifying harmful reactive oxygen species, and reducing oxidative stress.^24–26,39^. Unlike other studies, our randomized trial did not show a hepatoprotective effect. Various studies have reported NAC’s hepatoprotective effects against DS-TB-induced liver toxicity. In a study by Baniasadi et al. 2011, 60 patients with an average age of 60 or older and newly diagnosed TB were randomly assigned to two groups. 28 patients received oral NAC 600 mg twice daily in addition to drug-susceptible TB (DS-TB) treatment, while the 32 patients in the control group received only DS-TB treatment. None of the 28 patients in the intervention group developed liver toxicity, whereas 17 out of 32 patients in the control group developed liver toxicity within two weeks of evaluation. Similarly, in a study by Ahmed and Rao et al. 2020, 162 patients aged 18 to 60 with TB were divided into two groups per treatment arm. Half of the patients (81/162) in the control group received DS-TB treatment without NAC, while the other half (81/162) in the intervention group received both NAC 600 mg twice daily with DS-TB for eight weeks. In the control group, 17 out of 81 patients developed liver injury two weeks after the initiation of DS-TB treatment, compared to only 1 out of 81 patients in the interventional group.^24^ As a result, each study concluded by recommending NAC in combination with drug-susceptible TB treatment to reduce liver toxicity, a practice that has not been widely adopted. We found less liver injury and so could not distinguish a benefit of NAC, and as a consequence, were likely underpowered to do so. We suspect there are fewer events of liver toxicity given that rifampin and isoniazid were not part of this study regimen, as is the case with drug regimens, which may accelerate liver injury that is not observed in most RR/MDR-TB regimens.^10,40,41^ Our study population also had fewer women than in other trials of new RR/MDR-TB regimens. It has been suggested that women were more prone to liver injury compared to males in other studies of TB treatment.^41^ Other studies of more undernourished populations than our current study also suggest additional risk of TB treatment-induced liver injury.^42^ To gain more conclusive results, a larger trial involving patients with RR/MDR-TB may be necessary to observe any potential protective effects on the liver from NAC.

While we tested two dosing regimens of NAC, the total bioavailability may have needed to be increased to produce the maximum impact.^43^Usual oral dosing of NAC ranges 600-1200 mg up to 3 times daily, or higher dosage in psychiatric illnesses. Intravenous (IV) NAC reaches a maximum concentration of 500 mg/L within 15 minutes of 150 mg/kg administration, and IV dosing ranges from 50 mg/kg to 150 mg/kg as used in acetaminophen overdose, neurodegenerative diseases, post-cardiac surgery, and contrast-induced acute kidney injury.^16^ In our study, we used effervescent oral NAC at doses 900 mg daily and 1800 mg split over twice daily administration; these dosages were within frequently used oral dosages for anti-inflammatory and anti-oxidative effects. In Green et al., effervescent NAC demonstrated bioequivalence to oral NAC, and we chose effervescent because of its feasibility compared to IV NAC and tolerability compared to the oral pill form of NAC.^44^ Nevertheless, this is the first trial to our knowledge of long-term (24-week) NAC administration, which predicted that the intracellular benefits of NAC may accumulate over such a long time. Future studies could consider a higher dose of oral or IV formulations administered in the first week of therapy that could be transitioned to a more conventional oral dosing regimen concurrent with the remainder of TB treatment.

Following the TB treatment interval, NAC carries a potential as an approach to preventing post-TB lung disease (PTLD) - a debilitating respiratory condition estimated to impact half of TB survivors after cure.^45^ One postulated pathophysiology of PTLD is an imbalance in inflammatory and oxidative responses following TB disease and treatment.^45–47^ Given NAC’s role in immune modulation and replenishing glutathione, breaking disulfide bonds, restoring thiol stores, and enhancing anti-TB drug anti-mycobacterial effects, NAC may prevent the adverse remodeling and fibrotic response to tissue injury during and following TB treatment.^17^NAC is currently used to improve persistent chronic obstructive pulmonary disease exacerbations^48^and has demonstrated efficacy in increasing survival in the primary composite end point of death, hospitalization, or decline in lung function for individuals with idiopathic pulmonary fibrosis associated with rs3750920 (*TOLLIP*) TT genotype^49^ Further plans for the randomized groups in this current trial include a follow-up to determine the long-term benefits of NAC; including preventing signs and symptoms of PTLD.

Several limitations affected the generalizability and interpretation of the trial. The study was not designed to evaluate optimal NAC concentrations on glutathione levels. While two different dosing regimens were tested, the primary outcome was clinical and laboratory-measured AEs, and the relationship between specific AE development and glutathione level was unknown. Furthermore, while consistent trends in the prevention of AEs and SAEs from renal toxicity and the time to renal toxicity were observed in those patients receiving NAC, secondary assessments were not performed of cystatin-c or other non-enzymatic measurements of kidney function to determine if enzymatic interference could have been observed with high NAC exposures.^50,51^

In conclusion, this is the first randomized study on the effects of NAC in reducing AEs associated with RR/MDR-TB treatment. The findings show that NAC may have the potential drug to reduce renal toxicity, an event that frequently leads to hospitalization, treatment interruption, or regimen change. A conclusive and larger scale clinical trial should test the renoprotective effect of NAC in the RR/MDR-TB population while also incorporating a dose-finding lead-in of NAC plasma exposure and glutathione levels, trial outcomes to include non-enzymatic assessment of kidney function along with conventional creatinine measurement, and longer follow-up to determine the impact on lung health post TB cure.

## Supporting information

Comparing incidence of adverse events between combined NAC group and control group

Definitions of Adverse events

Liver injury time to event curve

Anemia time to event curve

## Data Availability

All data produced in the present study are available upon reasonable request to the authors
All data produced in the present work are contained in the manuscript
All data produced are available online at

## Acknowledgement

We want to express our deepest thanks to the participants, whose valuable contributions and willingness to engage were indispensable to generating meaningful and robust findings. We are also grateful to the Ministry of Health for their essential guidance and resources, which significantly contributed to the study’s successful completion. Furthermore, we extend our appreciation to the Kibong’oto Infectious Diseases Hospital healthcare workers and sponsorship that ensured critical support and the successful completion of this study. We appreciate Amelia Dana Badipour’s assistance with manuscript preparation.

## References

1. Global Tuberculosis Report 2023. Accessed May 6, 2024. https://www.who.int/teams/global-tuberculosis-programme/tb-reports/global-tuberculosis-report-2023

2. WHO announces updated definitions of extensively drug-resistant tuberculosis. Accessed December 14, 2023. https://www.who.int/news/item/27-01-2021-who-announces-updated-definitions-of-extensively-drug-resistant-tuberculosis

3. Ormerod LP. Multidrug-resistant tuberculosis (MDR-TB): epidemiology, prevention and treatment. British Medical Bulletin. 2005;73–74(1):17-24. doi:10.1093/bmb/ldh047

4. WHO Consolidated Guidelines on Tuberculosis: Module 4: Treatment - Drug-Resistant Tuberculosis Treatment, 2022 Update. World Health Organization; 2022. Accessed November 16, 2023. http://www.ncbi.nlm.nih.gov/books/NBK588564/

5. Jang JG, Chung JH. Diagnosis and treatment of multidrug-resistant tuberculosis. Yeungnam Univ J Med. 2020;37(4):277–285. doi:10.12701/yujm.2020.00626

6. Kushemererwa O, Nuwagira E, Kiptoo J, Yadesa TM. Adverse drug reactions and associated factors in multidrug-resistant tuberculosis: A retrospective review of patient medical records at Mbarara Regional Referral Hospital, Uganda. SAGE Open Med. 2023;11:20503121231171350. doi:10.1177/20503121231171350

7. Coleman JJ, Pontefract SK. Adverse drug reactions. Clin Med (Lond). 2016;16(5):481-485. doi:10.7861/clinmedicine.16-5-481

8. Nunn Andrew J., Phillips Patrick P.J., Meredith Sarah K., et al. A Trial of a Shorter Regimen for Rifampin-Resistant Tuberculosis. New England Journal of Medicine. 2019;380(13):1201–1213. doi:10.1056/NEJMoa1811867

9. Conradie Francesca, Bagdasaryan Tatevik R., Borisov Sergey, et al. Bedaquiline– Pretomanid–Linezolid Regimens for Drug-Resistant Tuberculosis. New England Journal of Medicine. 2022;387(9):810–823. doi:10.1056/NEJMoa2119430

10. Bezu H, Seifu D, Yimer G, Mebrhatu T. Prevalence and Risk Factors of Adverse Drug Reactions Associated Multidrug Resistant Tuberculosis Treatments in Selected Treatment Centers in Addis Ababa Ethiopia. Journal of Tuberculosis Research. 2014;02:144–154. doi:10.4236/jtr.2014.23018

11. TB Prevalence in Tanzania | National Tuberculosis & Leprosy Programme. Accessed November 15, 2023. https://ntlp.go.tz/tuberculosis/tb-prevalence-in-tanzania/

12. Mpagama SG, Mvungi HC, Mbelele PM, et al. Protocol for a feasibility randomized controlled trial to evaluate the efficacy, safety and tolerability of N-acetylcysteine in reducing adverse drug reactions among adults treated for multidrug-resistant tuberculosis in Tanzania. Pilot Feasibility Stud. 2023;9:55. doi:10.1186/s40814-023-01281-7

13. Karimi P, Guantai A, Kigondu C, Ogaro T. Adverse Drug Reactions Among patients Being Treated For Multi-Drug Resistan Tuberculosis in Nairobi City County Helath Facilities. psk.ke. Published online 2017.

14. Lan Z, Ahmad N, Baghaei P, et al. Drug-associated adverse events in the treatment of multidrug-resistant tuberculosis: an individual patient data meta-analysis. Lancet Respir Med. 2020;8(4):383–394. doi:10.1016/S2213-2600(20)30047-3

15. Prasad R, Singh A, Gupta N. Adverse Drug Reactions with First-Line and Second-Line Drugs in Treatment of Tuberculosis. Annals of the National Academy of Medical Sciences (India). 2021;57(01):15–35. doi:10.1055/s-0040-1722535

16. Tenório MC dos S, Graciliano NG, Moura FA, de Oliveira ACM, Goulart MOF. N-Acetylcysteine (NAC): Impacts on Human Health. Antioxidants (Basel). 2021;10(6):967. doi:10.3390/antiox10060967

17. Mapamba DA, Sauli E, Mrema L, et al. Impact of N-Acetyl Cysteine (NAC) on Tuberculosis (TB) Patients—A Systematic Review. Antioxidants (Basel). 2022;11(11):2298. doi:10.3390/antiox11112298

18. Ejigu DA, Abay SM. N-Acetyl Cysteine as an Adjunct in the Treatment of Tuberculosis. Tuberc Res Treat. 2020;2020:5907839. doi:10.1155/2020/5907839

19. Mahakalkar SM, Nagrale D, Gaur S, Urade C, Murhar B, Turankar A. N-acetylcysteine as an add-on to Directly Observed Therapy Short-I therapy in fresh pulmonary tuberculosis patients: A randomized, placebo-controlled, double-blinded study. Perspect Clin Res. 2017;8(3):132–136. doi:10.4103/2229-3485.210450

20. Safe IP, Lacerda MVG, Printes VS, et al. Safety and efficacy of N-acetylcysteine in hospitalized patients with HIV-associated tuberculosis: An open-label, randomized, phase II trial (RIPENACTB Study). PLOS ONE. 2020;15(6):e0235381. doi:10.1371/journal.pone.0235381

21. Huang JW, Lahey B, Clarkin OJ, et al. A Systematic Review of the Effect of N-Acetylcysteine on Serum Creatinine and Cystatin C Measurements. Kidney International Reports. 2021;6(2):396–403. doi:10.1016/j.ekir.2020.11.018

22. Guo Z, Liu J, Lei L, et al. Effect of N-acetylcysteine on prevention of contrast-associated acute kidney injury in patients with STEMI undergoing primary percutaneous coronary intervention: a systematic review and meta-analysis of randomised controlled trials. BMJ Open. 2020;10(10):e039009. doi:10.1136/bmjopen-2020-039009

23. Xie W, Liang X, Lin Z, Liu M, Ling Z. Latest Clinical Evidence About Effect of Acetylcysteine on Preventing Contrast-Induced Nephropathy in Patients Undergoing Angiography: A Meta-Analysis. Angiology. 2021;72(2):105–121. doi:10.1177/0003319720950162

24. Ahmed S, Rao NA. Role of N-acetylcysteine(NAC) in preventing development of anti tuberculosis therapy(ATT) induced liver injury in pulmonary tuberculosis(PTB) patients, a simple randomized single blind clinical trial. European Respiratory Journal. 2020;56(suppl 64). doi:10.1183/13993003.congress-2020.5301

25. Sanabria-Cabrera J, Tabbai S, Niu H, et al. N-Acetylcysteine for the Management of Non-Acetaminophen Drug-Induced Liver Injury in Adults: A Systematic Review. Frontiers in Pharmacology. 2022;13. Accessed November 20, 2023. https://www.frontiersin.org/articles/10.3389/fphar.2022.876868

26. Sukumaran D, Usharani P, Paramjyothi GK, Subbalaxmi MVS, Sireesha K, Abid Ali M. A study to evaluate the hepatoprotective effect of N-acetylcysteine on anti tuberculosis drug induced hepatotoxicity and quality of life. Indian Journal of Tuberculosis. 2023;70(3):303–310. doi:10.1016/j.ijtb.2022.05.012

27. Kalolo A, Lalashowi J, Pamba D, et al. Implementation of the ‘Removed Injectable modified Short-course regimens for EXpert Multidrug Resistant Tuberculosis’ (RISE study) in Tanzania: a protocol for a mixed-methods process evaluation. BMJ Open. 2022;12(5):e054434. doi:10.1136/bmjopen-2021-054434

28. Santana-Santos E, Gowdak LHW, Gaiotto FA, et al. High Dose of N-Acetylcystein Prevents Acute Kidney Injury in Chronic Kidney Disease Patients Undergoing Myocardial Revascularization. The Annals of Thoracic Surgery. 2014;97(5):1617–1623. doi:10.1016/j.athoracsur.2014.01.056

29. Javaherforooshzadeh F, Shaker Z, Rashidi M, Akhondzadeh R, Hayati F. The effect of N-acetyl cysteine injection on renal function after coronary artery bypass graft surgery: a randomized double blind clinical trial. Journal of Cardiothoracic Surgery. 2021;16(1):161. doi:10.1186/s13019-021-01550-7

30. Modarresi A, Ziaie S, Salamzadeh J, et al. Study of The Effects of N-Acetylcysteine on Oxidative Stress Status of Patients on Maintenance-Hemodialysis Undergoing Cadaveric Kidney Transplantation. Iran J Pharm Res. 2017;16(4):1631–1638.

31. Cepaityte D, Leivaditis K, Varouktsi G, Roumeliotis A, Roumeliotis S, Liakopoulos V. N-Acetylcysteine: more than preventing contrast-induced nephropathy in uremic patients—focus on the antioxidant and anti-inflammatory properties. Int Urol Nephrol. 2023;55(6):1481–1492. doi:10.1007/s11255-022-03455-3

32. Efrati S, Dishy V, Averbukh M, et al. The effect of *N*-acetylcysteine on renal function, nitric oxide, and oxidative stress after angiography. Kidney International. 2003;64(6):2182–2187. doi:10.1046/j.1523-1755.2003.00322.x

33. Tan YK, Luo H, Kang GS, Teoh KL, Kofidis T. N-Acetylcysteine’s Renoprotective Effect in Cardiac Surgery: A Systematic Review and Meta-Analysis. Ann Thorac Cardiovasc Surg. 2022;28(2):138–145. doi:10.5761/atcs.oa.21-00132

34. Nyang’wa Bern-Thomas, Berry Catherine, Kazounis Emil, et al. A 24-Week, All-Oral Regimen for Rifampin-Resistant Tuberculosis. New England Journal of Medicine. 2022;387(25):2331–2343. doi:10.1056/NEJMoa2117166

35. Hussain SW, Qadeer A, Munawar K, et al. Determining the Incidence of Acute Kidney Injury Using the RIFLE Criteria in the Medical Intensive Care Unit in a Tertiary Care Hospital Setting in Pakistan. Cureus. 11(2):e4071. doi:10.7759/cureus.4071

36. Yaqub S, Hashmi S, Kazmi MK, Aziz Ali A, Dawood T, Sharif H. A Comparison of AKIN, KDIGO, and RIFLE Definitions to Diagnose Acute Kidney Injury and Predict the Outcomes after Cardiac Surgery in a South Asian Cohort. Cardiorenal Medicine. 2022;12(1):29–38. doi:10.1159/000523828

37. Acute kidney injury - ClinicalKey. Accessed June 29, 2024. https://www.clinicalkey.com/#!/content/playContent/1-s2.0-S0140673619325632?returnurl=https:%2F%2Flinkinghub.elsevier.com%2Fretrieve%2Fpii%2FS0140673619325632%3Fshowall%3Dtrue&referrer=https:%2F%2Fpubmed.ncbi.nlm.nih.gov%2F

38. Amaral EP, Conceição EL, Costa DL, et al. N-acetyl-cysteine exhibits potent anti-mycobacterial activity in addition to its known anti-oxidative functions. BMC Microbiology. 2016;16(1):251. doi:10.1186/s12866-016-0872-7

39. Baniasadi S, Eftekhari P, Tabarsi P, et al. Protective effect of N-acetylcysteine on antituberculosis drug-induced hepatotoxicity. Eur J Gastroenterol Hepatol. 2010;22(10):1235–1238. doi:10.1097/MEG.0b013e32833aa11b

40. Yew WW, Leung CC. Antituberculosis drugs and hepatotoxicity. Respirology. 2006;11(6):699-707. doi:10.1111/j.1440-1843.2006.00941.x

41. Zhao H, Wang Y, Zhang T, Wang Q, Xie W. Drug-Induced Liver Injury from Anti-Tuberculosis Treatment: A Retrospective Cohort Study. Med Sci Monit. 2020;26:e920350–1-e920350-8. doi:10.12659/MSM.920350

42. Warmelink I, Hacken NH ten, Werf TS van der, Altena R van. Weight loss during tuberculosis treatment is an important risk factor for drug-induced hepatotoxicity. British Journal of Nutrition. 2011;105(3):400–408. doi:10.1017/S0007114510003636

43. Guerini M, Condrò G, Friuli V, Maggi L, Perugini P. N-acetylcysteine (NAC) and Its Role in Clinical Practice Management of Cystic Fibrosis (CF): A Review. Pharmaceuticals (Basel). 2022;15(2):217. doi:10.3390/ph15020217

44. Greene SC, Noonan PK, Sanabria C, Peacock WF. Effervescent N-Acetylcysteine Tablets versus Oral Solution N-Acetylcysteine in Fasting Healthy Adults: An Open-Label, Randomized, Single-Dose, Crossover, Relative Bioavailability Study. Curr Ther Res Clin Exp. 2016;83:1–7. doi:10.1016/j.curtheres.2016.06.001

45. Singh S, Allwood BW, Chiyaka TL, et al. Immunologic and imaging signatures in post tuberculosis lung disease. Tuberculosis. 2022;136:102244. doi:10.1016/j.tube.2022.102244

46. Ravimohan S, Kornfeld H, Weissman D, Bisson GP. Tuberculosis and lung damage: from epidemiology to pathophysiology. Eur Respir Rev. 2018;27(147):170077. doi:10.1183/16000617.0077-2017

47. Malefane L, Maarman G. Post-tuberculosis lung disease and inflammatory role players: can we characterise the myriad inflammatory pathways involved to gain a better understanding? Chemico-Biological Interactions. 2024;387:110817. doi:10.1016/j.cbi.2023.110817

48. Calverley P, Rogliani P, Papi A. Safety of N-Acetylcysteine at High Doses in Chronic Respiratory Diseases: A Review. Drug Saf. 2021;44(3):273–290. doi:10.1007/s40264-020-01026-y

49. Oldham JM, Ma SF, Martinez FJ, et al. TOLLIP, MUC5B, and the Response to N-Acetylcysteine among Individuals with Idiopathic Pulmonary Fibrosis. Am J Respir Crit Care Med. 2015;192(12):1475–1482. doi:10.1164/rccm.201505-1010OC

50. Magner K, Ilin JV, Clark EG, Kong JWY, Davis A, Hiremath S. Meta-analytic Techniques to Assess the Association Between N-acetylcysteine and Acute Kidney Injury After Contrast Administration: A Systematic Review and Meta-analysis. JAMA Network Open. 2022;5(7):e2220671. doi:10.1001/jamanetworkopen.2022.20671

51. McCudden C, Clark EG, Akbari A, Kong J, Kanji S, Hiremath S. N-Acetylcysteine Interference With Creatinine Measurement: An In Vitro Analysis. Kidney International Reports. 2021;6(7):1973–1976. doi:10.1016/j.ekir.2021.04.006

