## Supplementary material for "N-acetylcysteine to reduce kidney and liver injury associated with drug-resistant tuberculosis treatment": Comparing incidence of adverse events between combined NAC group and control group

*Supplemental Table 1: Comparing the incidence of adverse events in the combined*

*N-acetylcysteine vs standard treatment group (N = 66)*

| systems | TAE in the standard treatment group | Total patients with at least one event in the standard treatment group  (N=22) | TAE in combined  NAC group | Total patients with at least one event in a combined  NAC group  (N = 44) | TAE in all patients across | Total patients with at least one AE across (N=66) | P-value |
| --- | --- | --- | --- | --- | --- | --- | --- |
| Nervous system | **0** | 0 | 2 | 2 (4.5 %) | 2 | 2 | 0.549 |
| Visual | **0** | 0 | 2 | 2 (4.5%) | 2 | 2 | 0.549 |
| Endocrine | **2** | 2 (9) % | 1 | 1 (2.3%) | 3 | 3 | 0.256 |
| Gastro  intestinal tract | **5** | 3 (14%) | 11 | 7 (16%) | 20 | 10 | 0.281 |
| Hepatic | **2** | 2 (9%) | 4 | 4 (9%) | 6 | 6 | 1.000 |
| Renal | **16** | 10  (45%) | 17 | 10 (22%) | 33 | 20 | 0.058 |
| Muscular skeletal | **15** | 6 (27%) | 43 | 13 (30%) | 58 | 19 | 0.442 |
| Skin | **3** | 3 (14%) | 1 | 1 (2.3%) | 4 | 3 | 0.104 |
| Hematology | **9** | 7  (32%) | 21 | 16  (36%) | 30 | 23 | 0.715 |

Note: *N-acetylcysteine (NAC), total adverse event (TAE),* Analysis with Fisher exact test/ꭓ^2^
