## Supplementary material for "N-acetylcysteine to reduce kidney and liver injury associated with drug-resistant tuberculosis treatment": Definitions of Adverse events

*Table 3: Definitions of Adverse events used in the study*

| Adverse event category | Definition |
| --- | --- |
| Cardiovascular  Dermatological  Central nervous system  Ocular  Gastrointestinal  Endocrine/metabolic changes  Musculoskeletal  Renal    Hepatic  Hematology | *Palpitations or QTc prolongation >/ 500ms on EKG*  *Report of skin rash, pruritis, color changes*  *Psychosis reported by a psychiatrist, seizures witnessed or reported by the patient, anxiety, depression*  *The patient reported visual changes*  *Nausea, vomiting, abdominal pain, diarrhea, constipation*  *Measurement of TSH 2.2-4.2µIU/ml, serum calcium 0.5-2.55 mmol/l,potassium 3.6-5.5 mmol/l*  *Joint pain, swelling, back pain, elevated uric acid, arthralgia*  *RIFLE -*  *mild – Serum creatinine level > 1.5 times baseline*  *moderate - serum creatinine level > 2 times baseline*  *severe- serum creatinine level >3 times above baseline, life-threatening: dialysis, death*  *Definition of Hepatitis using WHO’s recommendation:*  *1)ALT or AST five or more times ULN;2) ALT or AST three times the ULN with clinical manifestations; and or 3) ALT or AST three or more timed the ULN with a concomitant increase in bilirubin of >/1.5 times*    *The strongest confirmation of the diagnosis of DILI is > two-fold serum ALT elevation and discontinuation leading to a fall in ALT levels*    *Elevated transaminases/bilirubin*  *Grade 1-5, severe adverse events grade 3 and above*  *Alanine Transferase (ALT)*  *Grade 1 – 1.5- 3 times ULN with normal baseline: 1.5-3 times baseline, with abnormal baseline*  *Grade 2 - > three times ULN with normal baseline: >3 times baseline, with abnormal baseline*  *Grade 3 - > five times ULN with normal baseline: > 5 times baseline, with abnormal baseline*  *Grade 4 - > 20 times ULN with normal baseline: > 20 times baseline, with abnormal baseline*  *Aspartate aminotransferase (AST)*  *Grade 1 – 3 times ULN with normal baseline: 3 times baseline, with abnormal baseline*  *Grade 2 - > three times ULN with normal baseline: >3 times baseline, with abnormal baseline*  *Grade 3 - > five times ULN with normal baseline: > 5 times baseline, with abnormal baseline*  *Grade 4 - > 20 times ULN with normal baseline: > 20 times baseline, with abnormal baseline*  *Bilirubin*  *Grade 1-4*  *Grade 1- 1.5 x ULN if the baseline was normal;1.0-1.5 x baseline if the baseline was abnormal.*  *Grade 2- > 1.5 x ULN if the baseline was normal; > 1.5 x baseline if the baseline was abnormal.*  *Grade 3- > 3 x ULN if the baseline was normal;> 3 x baseline if the baseline was abnormal.*  *Grade 4- >10 x ULN if the baseline was normal;> 10 x baseline if the baseline was abnormal.*  *A decrease in white cell count (leukopenia), platelets(thrombocytopenia), or hemoglobin (anemia) below the standard limit.*  *Grade 1-4; severe adverse event grade 3 and above*  *Anemia*  *Grade 1 < normal limits*  *Grade 2 < 10 g/dL*  *Grade 3 < 8 g/dL*  *Grade 4- life-threatening*  *Thrombocytopenia*  *Grade 1 < 150,000*  *Grade 2< 75,000*  *Grade 3< 50,000*  *Grade 4 < 25,000*  *Leukopenia*  *Grade 1 < 5000/mm^3^*  *Grade 2 < 3000/ mm^3^*  *Grade 3 < 2000/ mm^3^*  *Grade 4 < 1000/ mm^3^* |

Notes: Upper limit normal (ULN), aspartate transaminase (AST), alanine transaminase (ALT), risk, injury, failure, loss and end stage kidney disease (RIFLE), Drug induced liver injury (DILI).
