## Supplementary material for "N-acetylcysteine to reduce kidney and liver injury associated with drug-resistant tuberculosis treatment": Liver injury time to event curve

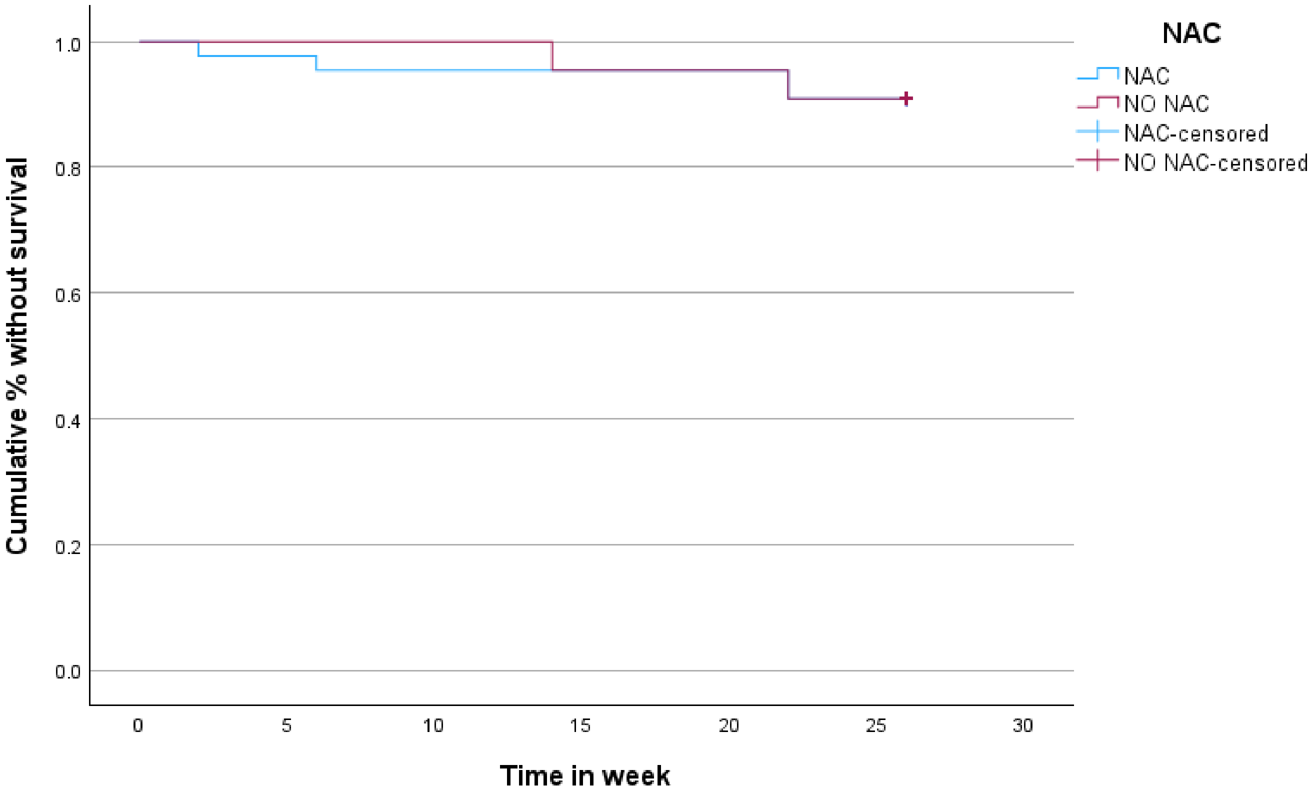


Supplemental Figure1: liver injury time to event liver injury curve through 26 weeks with standard treatment and NAC
