## Supplementary material for "N-acetylcysteine to reduce kidney and liver injury associated with drug-resistant tuberculosis treatment": Anemia time to event curve

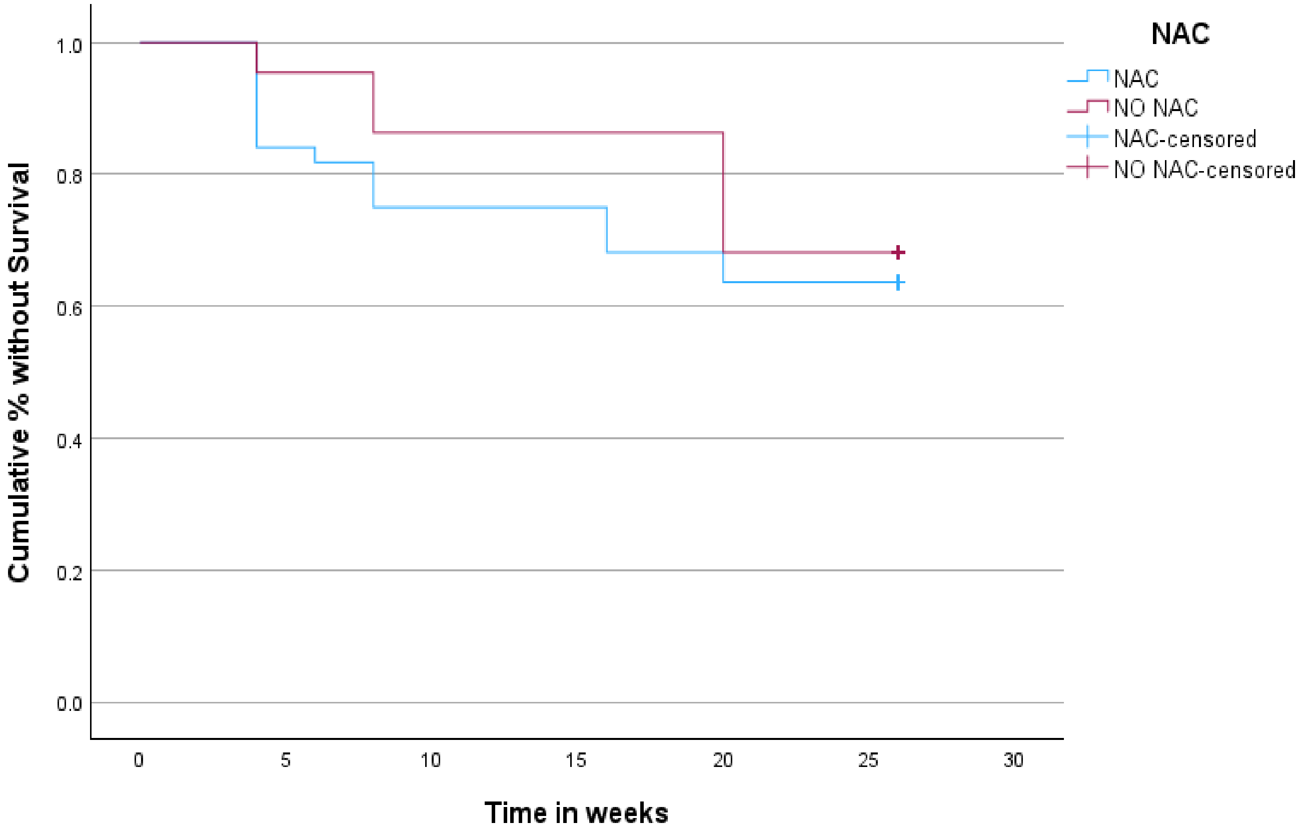


supplemental figure 2: anemia time to event curve through 24 weeks between standard treatment and NAC group.
